# Building an Analytical Framework for Tobacco-Related Misinformation on Social Media: An Exploratory Analysis with Generative AI Assistance

**DOI:** 10.1101/2025.05.20.25328002

**Authors:** Eileen Han, Miao Feng, Pamela Ling

## Abstract

**Background:** The propagation of tobacco-related misinformation significantly impacts public health, particularly affecting people with less access to reliable information sources (such as those with lower education), who may also su affer disproportionate tobacco-related morbidity and mortality. This study analyzed a dataset from Twitter to identify the characteristics of tobacco-related misinformation, with the goal of creating a framework for its identification, categorization, and validation.

**Methods:** A collection of 3.4 million tweets related to tobacco and nicotine was refined to 842,754 after removing irrelevant and duplicate posts. LDA topic modeling identified six unique topics, from which two randomly selected samples of tweets were drawn to perform qualitative analysis and AI-assisted analysis to identify categories of tobacco misinformation.

**Results:** The identified tobacco-related misinformation was categorized by three dimensions (1) content, including safety and health effects, cessation, substance, and policy; (2) type of falsehood, which included fabrication and unsubstantiated claims, misrepresentations, and distortions; and (3) source, ranging from individuals and retail stores to advocacy groups and influencers.

A notable finding was the prevalence of policy-related discussions of tobacco misinformation on Twitter (X), highlighting this often-overlooked domain. The controversy over vaping has amplified pro-vaping voices on social media, with content frequently misinterpreting scientific findings, policies, and expert opinions, reflecting more nuanced and difficult to recognize falsehood in the misleading content.

**Conclusion:** This study offers a comprehensive framework for analyzing tobacco-related misinformation on social media, emphasizing key issues in policy debates and the presence of conspiracy narratives. This framework can inform the design of interventions for less informed populations and enhance data annotation for machine learning tasks.

## Background

Tobacco-related misinformation on social media can significantly impact public health by shaping false perceptions and beliefs. Misinformation may mislead users of tobacco products in decision-making and potentially harms the health of communities most impacted by tobacco product use and its health consequences, contributing to health disparities (1). Smoking is more concentrated among some groups of people, such as those with lower levels of income or education, who also have less access to reliable information (2), increasing the potential impact of misinformation on them. For example, misperceptions about the risks and benefits of e-cigarettes could affect decisions to switch or quit (3). With the tobacco industry’s continuing targeted marketing, inconclusive data on the risks and benefits of emerging novel products, and inequalities in accessing reliable and quality information, improving information literacy could mitigate disparities in tobacco-related health outcomes (4).

While health misinformation studies are abundant, as smoking and drug-related misinformation is prevalent on X (previously and herein: Twitter) (5), relatively few studies directly address tobacco-related misinformation on social media. Understanding the common characteristics of tobacco misinformation on social media could help public health researchers and experts design interventions for targeted populations. However, as new tobacco products rapidly emerge, misinformation related to these products also grows quickly, with potentially misleading marketing messages and social media discussions (1). It is challenging for evidence-based research and public health surveillance systems to keep up and address the potential risks of new and evolving products (6). A framework that captures common patterns and characteristics of existing tobacco misinformation and that can adapt to emerging trends can inform our understanding of misinformation and strategies to correct misunderstanding.

Previous studies on tobacco misinformation have focused on single issues, such as misunderstanding of nicotine, the health risks and benefits of e-cigarettes, or the relationship between vaping and COVID-19 (7–10). Some studies have tested selected message effects (11,12) and corrective messages (13). However, to our knowledge, no study provided ways to systematically organizes tobacco-related misinformation on social media. In the area of vaccines, researchers have developed typologies or taxonomies about anti-vaccine misinformation based on online content and design features (14,15). In this study, we drew on Twitter data, identified key components of tobacco-related misinformation with the assistance of generative AI tools, and explored ways to organize tobacco-related misinformation on social media to support further research into its characteristics and the development of tailored counter-messages.

## Methods

### Data Collection Rationale

The tobacco industry has been identified as a source of misinformation through misleading public relation posts on social media(16,17). Studies also show that while the tobacco industry’s social media accounts may seem compliant with marketing restrictions, such as setting age limits for content viewing and refraining from using words indicating health benefits, industry messages can share hashtags with public tweets produced by others about the same topic without such restrictions (18). Therefore, tobacco industry-produced content serves as a reasonable starting point for exploring the use of similar hashtags and keywords in a broader context. In this study, we started with an analysis of industry tweets to identify priority topics in recent years and used their top-prioritized hashtags and corresponding keywords (hashtags) to further inform our search for general public tweets. All tweets from two leading transnational tobacco companies, British American Tobacco and Philip Morris International, were collected using the Twitter application programming interface (API) from January 2020 to December 2022.

Figure 1 shows the development process for search queries. The top 10 hashtags the two tobacco companies used between 2020 and 2022 were identified in the dataset of industry tweets, and company-specific-hashtags (with company names or slogans in them) were excluded. Eight hashtags remained in the search query, and together with their corresponding keywords (Q1), they were used to search tweets in a larger database that collected public tobacco-related posts since 2015 using Twitter’s streaming API (19) (Figure 1). The search resulted in a total of 3,436,070 tweets dated from January 1, 2020, to December 31, 2022 (Public Tweets). Because these search keywords included hashtags used by tobacco companies are widely used in other contexts (e.g. #science and #innovation), the resulting public tweets contained many thematically irrelevant tweets . Therefore, an additional search of the public tweets was conducted with a few tobacco-related key terms that did not appear in the initial search but were commonly used in tobacco-related tweets (Q2), ensuring that the final set of tweets all contained tobacco-related terms (Figure 1). For example, while “e-cigarette” and related terms (“vape”, “e-cig”, “ecig”) were not seen in the industry’s top hashtags, they were still frequently used in public tweets that contain any keywords in Q1. By combining the search results from Q1 and Q2, 842,754 tweets (final set) were found after duplicates were removed. In other words, each of the resulting tweets in the final set contains any keywords in Q1 and any keywords in Q2.

**Figure 1.**
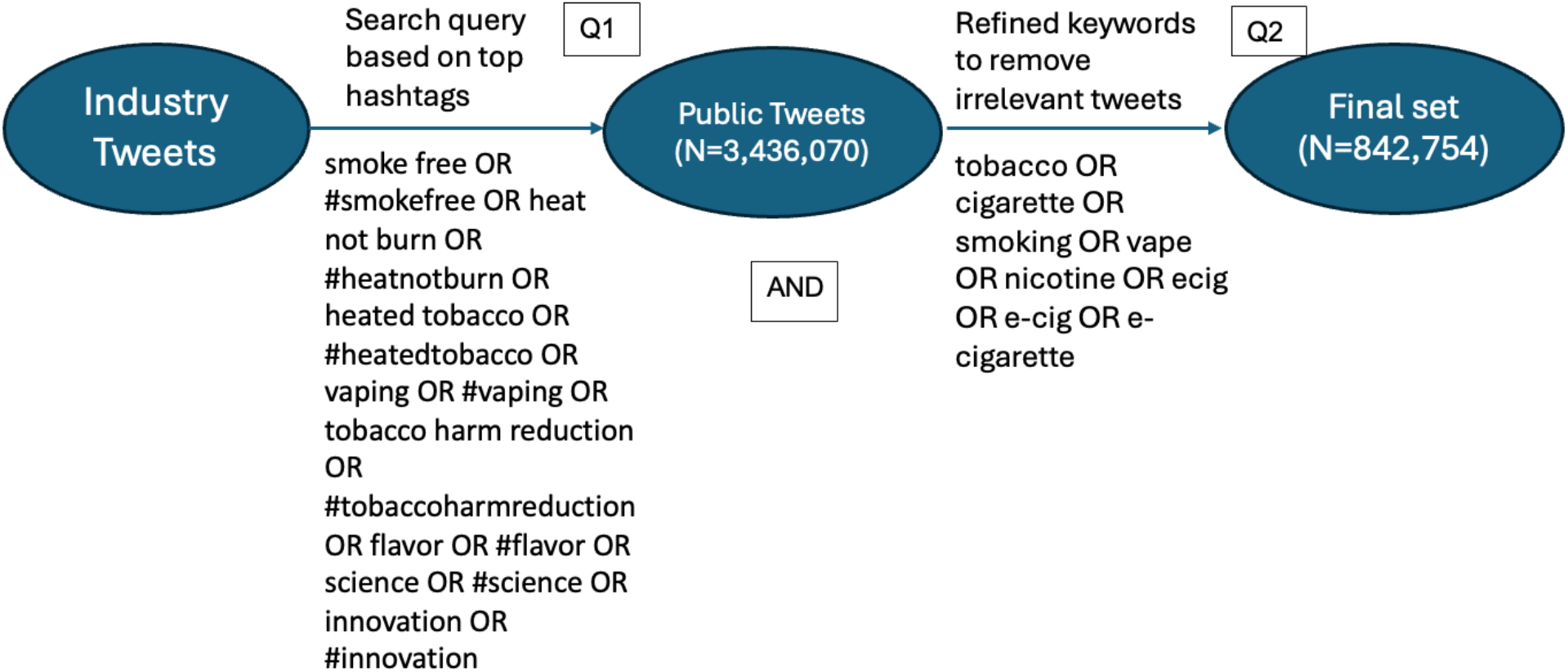
Data Collection and Processing

### Data Analysis

Topic modeling was performed using the LDA model (20) for this collection of 842,754 tweets. We selected the 6-topic model as it resulted in the highest coherence score among the number of topics ranging from 3 to 20. We reviewed a random selection of 100 tweets from each topic and named each topic based on the main themes present in the sample tweets. The 600 tweets served as our initial sample (S1) that all three researchers reviewed in the qualitative analysis to identify key components of the framework.

The qualitative analysis of the tweets involved three steps. First, we built our selection criteria by consulting experts and referring to guidelines from established health authorities (21–25). We curated a list of claims containing misinformation related to different aspects of tobacco products from the literature, including the safety of e-cigarettes, the addictiveness of nicotine, nicotine’s therapeutic functions, e-cigarette/smoking and COVID-19 association, and tobacco company self-promotion. (8,9,16,26) These claims were used as our “seed” claims (27). We also referred to a recently published scoping review of health misinformation to define key components of misinformation, such as being false, inaccurate, or incorrect, with or without harm intention, and having possible misleading effects (28).

Next, we reviewed each tweet in S1, identifying concepts that matched those mentioned in our seed claims. We then compared the main claim in the tweet with our guideline references and its similarity to the seed claims to determine if that the tweet should be categorized as misinformation. We also noted concepts that were not in the seed claims and created new categories, iteratively building the framework.

### Here is a redacted example

*RT @xxx: @xxx, @xxx, @xxx Did you know that 97% of people use vaping devices as a way to quit a dangerous habit? Vaping is considered the safest alternative to smoking traditional cigarettes. Let’s educate young people to make wise choices and avoid smoking! In my opinion, compassion and understanding of this topic are essential, which is why it’s clear that no one actually ‘smokes’ an e-cigarette*.

The key concepts we identified in this tweet were smoking cessation and the safety of vaping. Compared with the guidelines, the claim “97% of people use vaping devices as a way to quit a dangerous habit” was not supported because neither the CDC and WHO have affirmed the number or percentage of e-cigarette users who intend use for smoking cessation, or the effectiveness of vaping as a cessation tool.

After reviewing all tweets in the sample, researchers deliberated on their selections, through which we finalized a few selection standards, particularly on how tweets expressing personal experiences should be categorized, and how to categorize tweets that were addressing controversial topics.

When identifying misinformation, if a tweet was mainly an expression of personal opinion or testimonies, it did not necessarily qualify as misinformation. However, if that personal testimony or opinion implied generalizability, it was classified as misinformation. In this example, the phase “vaping is considered the safest alternative” would be identified as misinformation both because it is not accurate, (as CDC and WHO endorse approved pharmacotherapy or complete tobacco cessation as safer than use of e-cigarettes), and the phrase is presented as a generalizable true statement.

The topic of smoking cessation included a mixture of personal experiences and general discussions about the benefits of e-cigarettes as a cessation tool. We categorized all tweets suggesting vaping as an effective cessation tool for people in general as misinformation, following the WHO’s guidelines indicating insufficient evidence of the effectiveness of e-cigarette use for cessation at the population level. For instance, if someone tweeted about successfully quitting smoking with vaping, or stated ‘I think vaping is a more effective cessation tool than other methods,’ this was not classified as misinformation because it reflects a personal experience that could be true for that individual. In contrast, a tweet claiming, “I used e-cigarettes to quit smoking, and everyone should have access to vaping, the best cessation tool ever” would be classified as misinformation because it generalizes the effectiveness of vaping as cessation tool, a claim insufficiently supported by scientific evidence.

Similarly, we considered the nuances in claims for topics where there is scientific controversy or contentious discussion, such as the safety of vaping. We did not consider tweets arguing for the relative safety of vaping compared to cigarettes as misinformation, as this claim has gained endorsements from some scientific consensus groups, although these groups may have different views about whether e-cigarettes should be used as a cessation tool (29,30). However, any claim using absolute terms to discuss a nuanced topic like the safety and health of vaping was classified as misinformation. For example, a claim stating “vaping is totally safe” or “vaping is the best alternative to smoking” would be classified as misinformation.

Next, we compared this classification system to one produced by generative AI (ChatGPT 4.0). We prompted ChatGPT, a conversational AI system (31), to provide its standard for selecting tobacco-related misinformation as if it were a public health expert in tobacco control. Based on its current knowledge, following response was generated:

- *Claim that vaping or e-cigarette use is completely safe or significantly safer than traditional tobacco use without acknowledging the existing health risks;*
- *Promote tobacco products as a healthy alternative to smoking;*
- *Suggest that tobacco products have health benefits;*
- *Target younger audiences or non-smokers, encouraging them to start using tobacco products;*
- *Misrepresent the addictive nature of nicotine or tobacco*.

Compared with the human-generated standard, the AI standard did not mention cessation but focused on a more general “benefit”. The AI standards also used the term “tobacco products” rather than “e-cigarettes” or “vaping” in several statements, leaving uncertainty about whether the term “tobacco products” would include or exclude e-cigarettes. In general, the statements were very close to what we generally relied on for misinformation classification standards. It was not clear how the AI would classify a nuanced statement or take into account personal opinion as explained above.

We then asked AI to apply these standards to examine the 600-tweet sample and make selections and define categories of misinformation. The results were then validated by human researchers, and the tweets that were identified by AI and judged to correctly identify misinformation were incorporated in the final results.

Next, in order to test the ability of ChatGPT to help automate the misinformation selection and categorization process, we created a larger sample (S2, 1% of the dataset, N=8427), proportional to the sizes of each of the six topics from the topic modeling. Reading the sample tweets and seed claims, it generated selection standards similar to the above ones, and added two standards related to more recent events: 1) nicotine’s protective effects on COVID-19, and 2) EVALI is unrelated to vaping. It then produced a few categories that it considered as misinformation. These two additional standards could be resulted from seeing the presence of tweets discussing COVID-19 and EVALI. Finally, we asked ChatGPT to provide its own selection of tweets containing misinformation from the sample, and then validated its selection with human researchers. The validated tweets were also included in the final selection (Figure 2).

**Figure 2.**
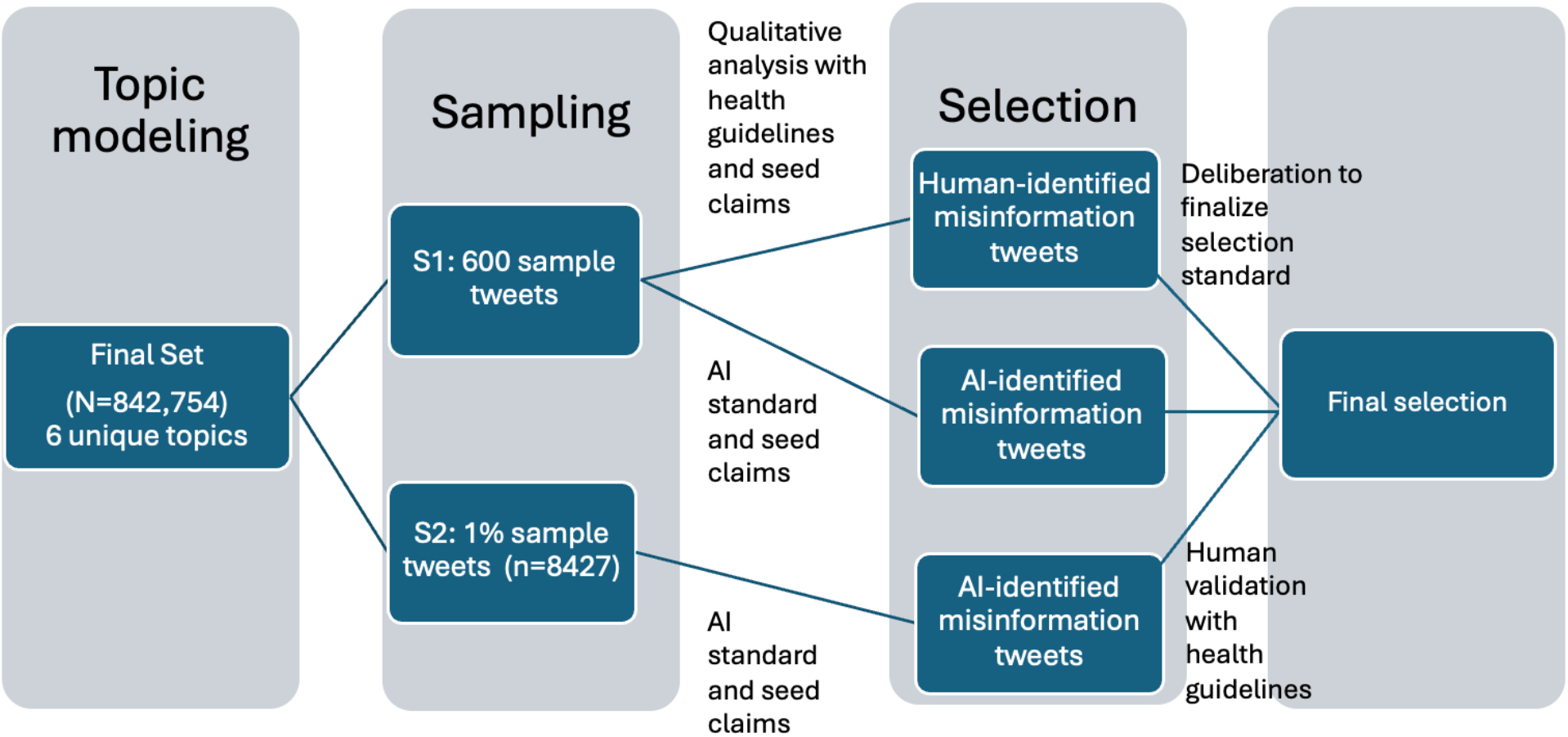
Data Analysis Process

**Figure 3.**
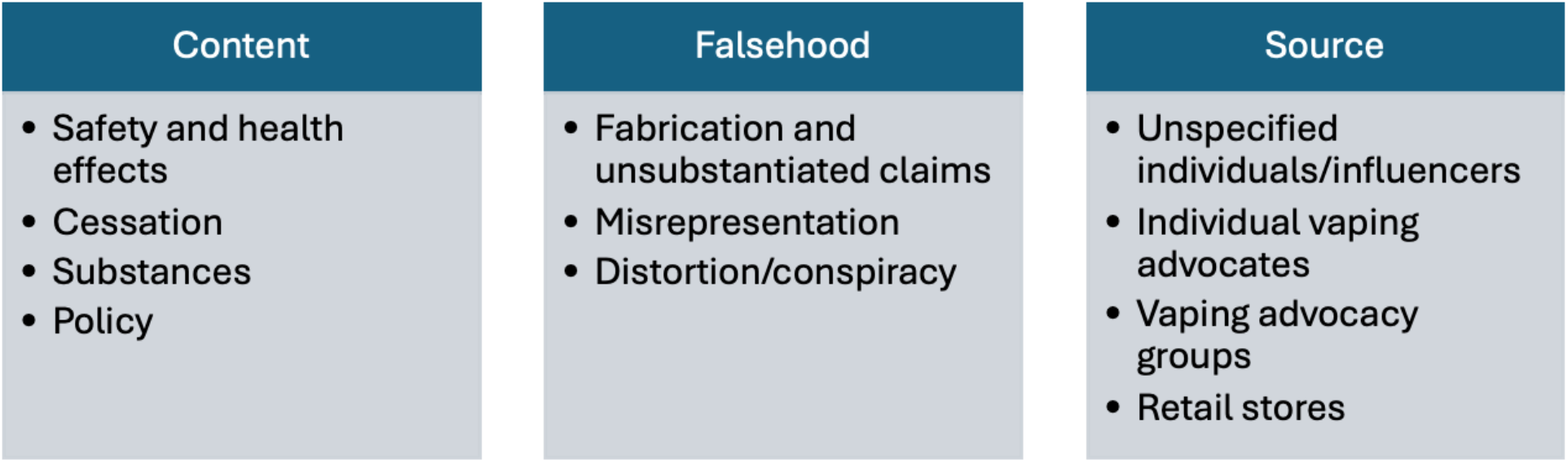
The Tobacco Misinformation Framework

## Results

Six unique topics resulted from topic modeling. These topics informed us the main themes of tweets in this large dataset. The topic names, topic sizes (number of tweets), and redacted examples are listed in Table 1. From each topic, we randomly selected 100 tweets for further analysis, making the 600-tweet analytical sample. Then, a larger sample of 8427 tweets was drawn, proportional to the size of each topic, for AI-assisted analysis.

**Table 1.**
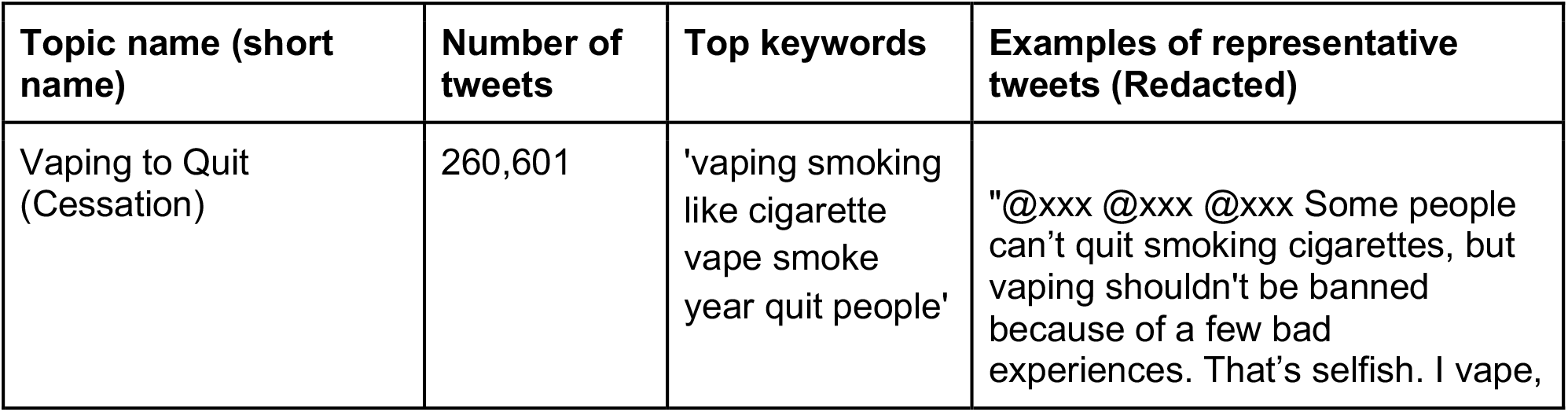
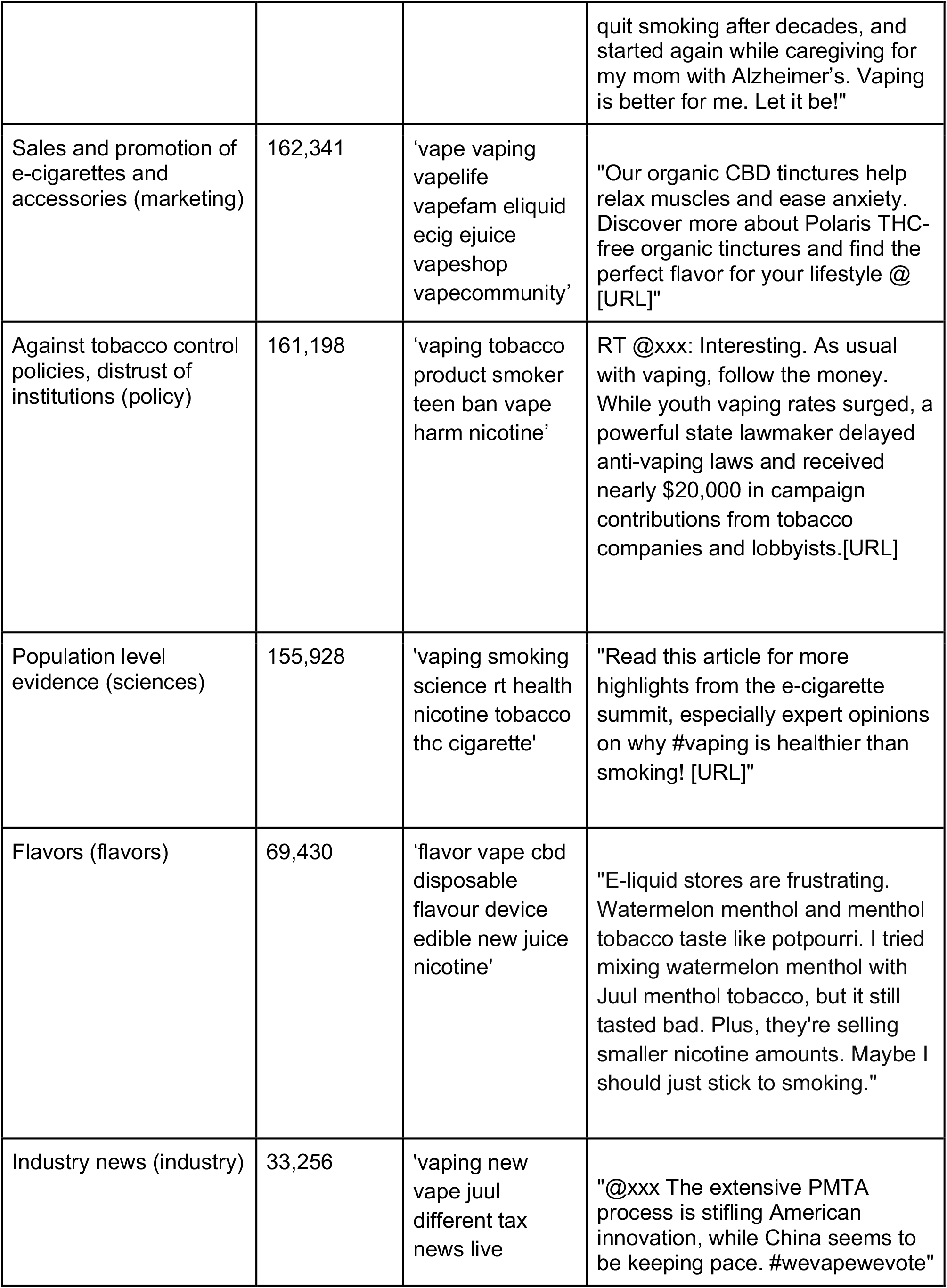
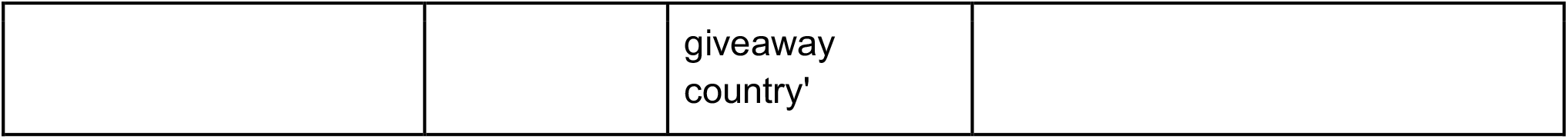
Results from Topic Modeling.

Out of the 600 sample tweets in S1, human coders identified 58 as containing misinformation, while the AI identified 21 tweets as misinformation, and eight of which were also identified by human (a subset of the 58), all containing pro-vaping content. The other 13 AI-identified tweets did not meet the health guidelines and researcher agreements and were thus excluded from the final selection. For the additional 8427 tweet-sample (S2), 557 tweets were identified using the AI standard, and 121 were validated by human coders applying the official health guidelines. Our final selection of tweets containing misinformation was 169, including the 58 human identified tweets in S1 and the 121 identified by AI and human validated.

Through analyzing the tweets that contained misinformation in the sample, we identified three key components to categorize tobacco-related misinformation: 1) content, 2) type of falsehood, and 3) source. Misinformation categories provided by AI include: Nicotine reduces COVID-19 risks, vaping is completely safe and risk free, EVALI is unrelated to vaping, nicotine is not or slightly addictive, economic and regulatory influences, and vaping is an effective cessation tool for everyone. Compared with human categorization, the AI categories were one dimensional, focusing on content, and did not differentiate the types of falsehood in those claims or consider sources because this dataset does not have user information directly accessible. AI also mistakenly identified tweets expressing relative safety of vaping as misinformation, possibly due to the tendency to classify the presence of any “safe” or “healthy” terms as misinformation.

### Content

The content dimension was mainly informed by the topic modeling results and the categories provided by AI. Within misinformation content we identified four major subtopics that were both identified by human and AI: safety and health effects, cessation, substances, and policy. (Table 2) Among the 179 selected misinformation tweets, 79 were about safety and health effects (mainly about vaping), making it the largest misinformation topic. Policy was also a common topic (49 tweets).

**Table 2.**
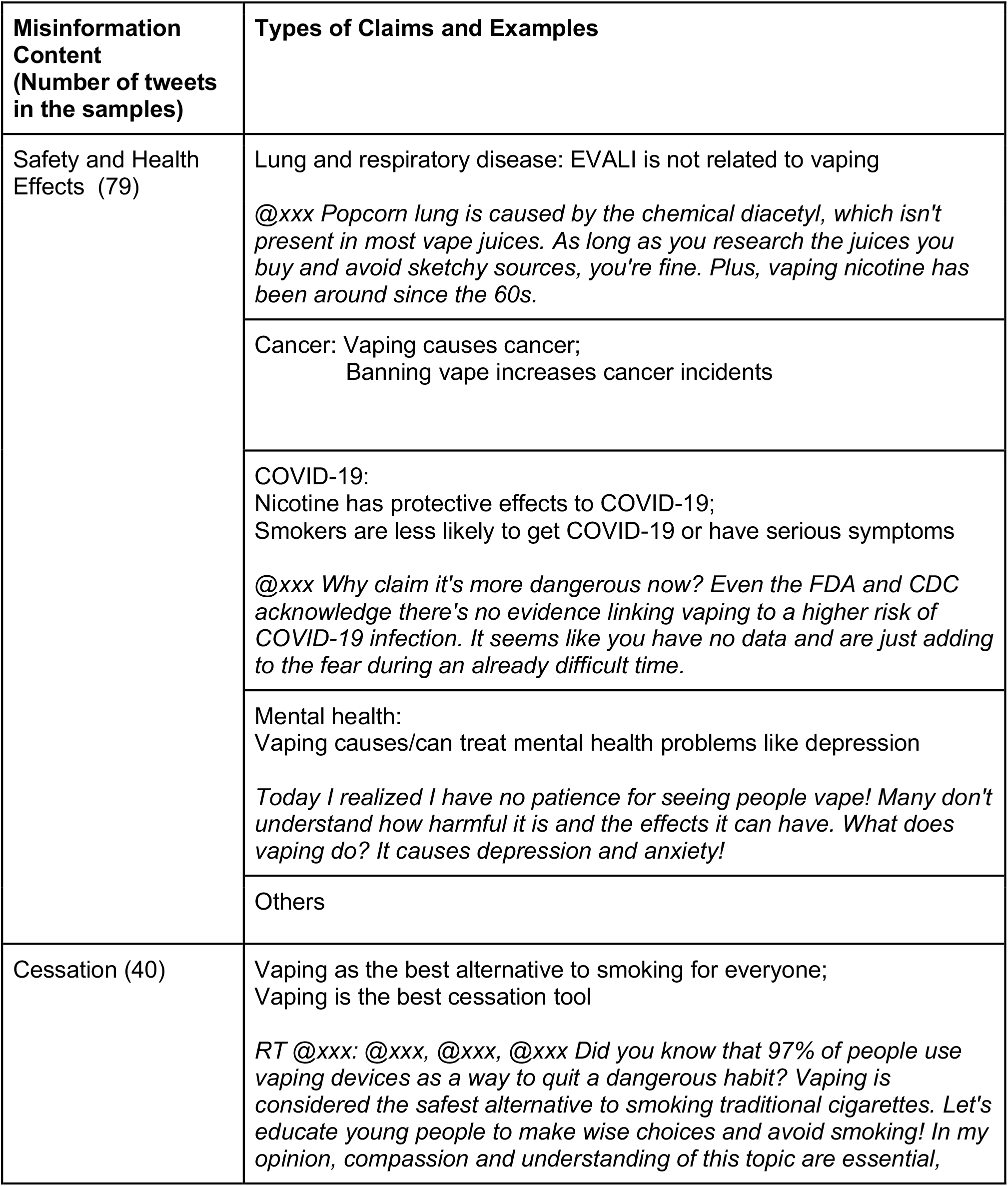
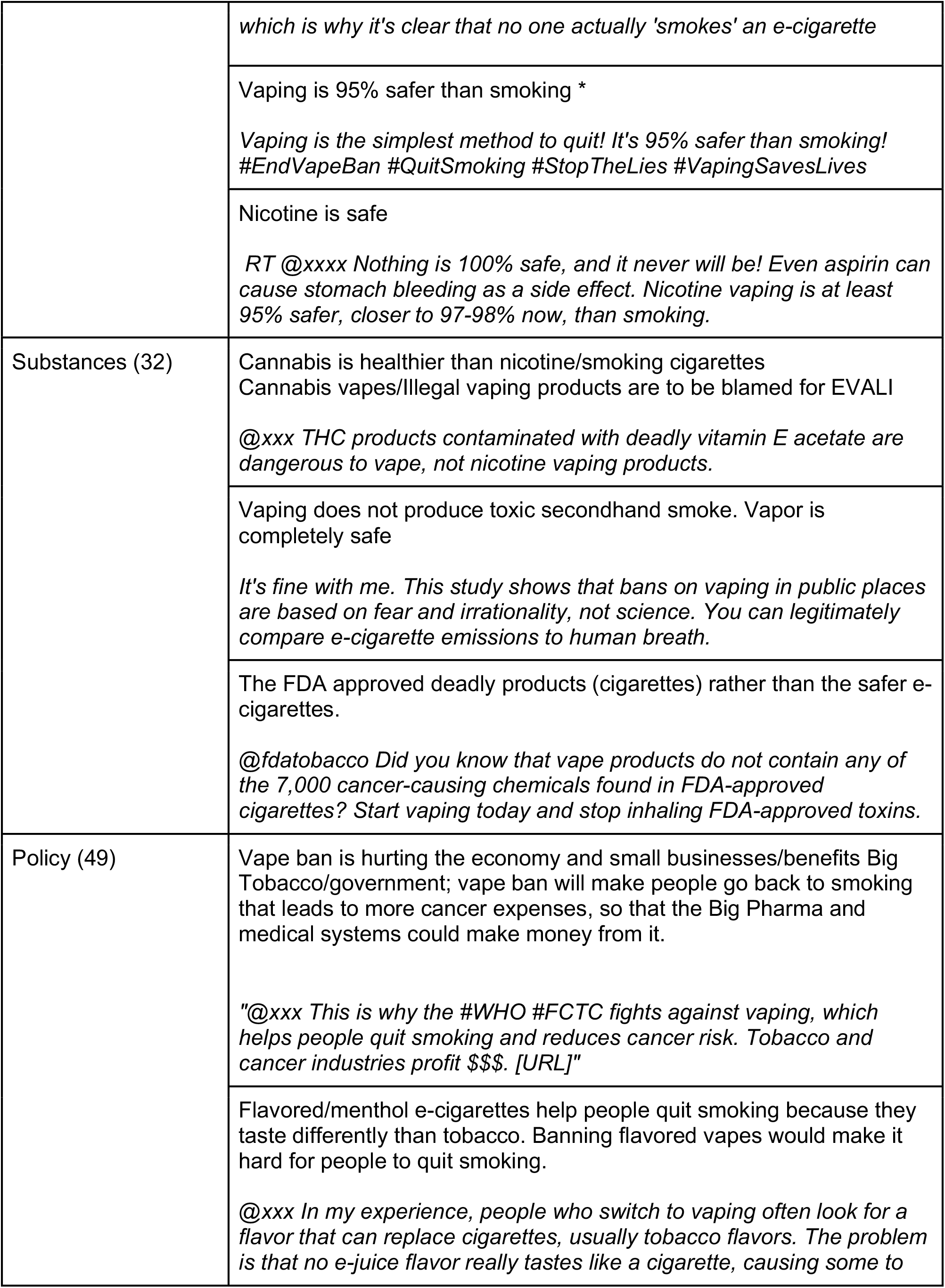
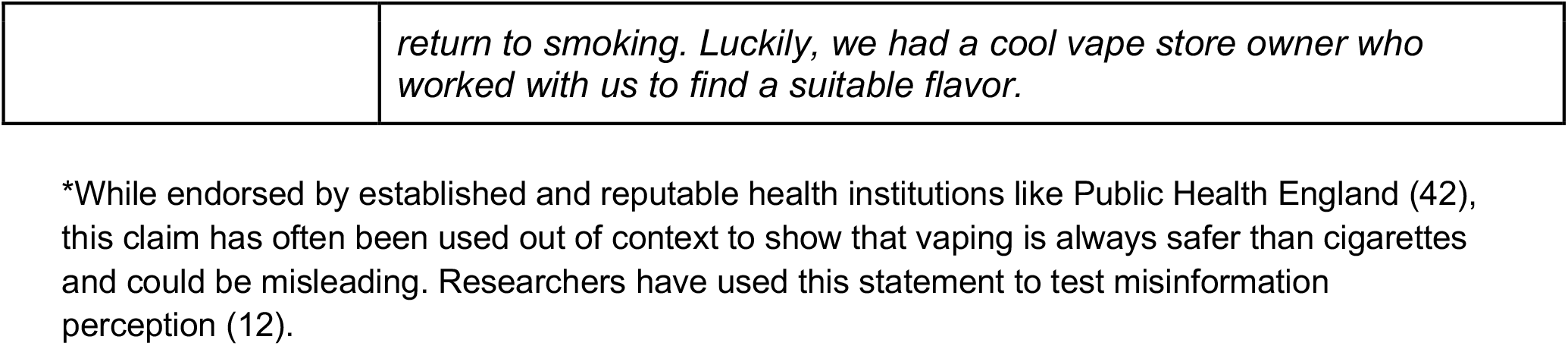
Misinformation content and examples.

### Types of Falsehood

The falsehood of the claims refers to the degree to which they deviate from reality or existing scientific consensus. Drawing on existing typology (14), we identified three types of falsehood: (a) fabrication and unsubstantiated claims, (b) misrepresentation, and (c) distortion/conspiracy. Among the 169 misinformation tweets, 30 were fabrication and unsubstantiated claims, 122 were misrepresentation, and 44 were distortion/conspiracy.

#### Fabrication and Unsubstantiated Claims

Fabrication and unsubstantiated claims include those with factual errors or those lacking scientific evidence. For example, the claim “Vaping reduces the risks of COVID-19” is a fabrication, as there is no scientific evidence to support it.

#### Misrepresentation

Misrepresentation involves claims that may be true or partially true but are inaccurately presented or interpreted. These claims can be misused (cited out of context), exaggerated (overstated), or employed with inappropriate analogies, leading to misleading conclusions. In our analysis, 36 of the 58 human-identified misinformation tweets fell into this category.

For instance, the claim “Vaping is completely safe and risk-free” is an exaggeration. Similarly, “Vaping is an effective cessation tool for me, so it should work for everyone” overstates its benefits, neglecting the specific conditions under which vaping can be an effective cessation tool, such as adult use only, complete switching from cigarettes, and the fact that it is never safe for youth or non-smokers (21). A notable pattern in our analysis was the misinterpretation of policies.

Consider the tweet: “Are you aware that vaping doesn’t contain any of the numerous carcinogenic chemicals found in traditional cigarettes that are approved by the FDA in every cigarette? Make the switch to vaping and avoid breathing in the harmful compounds that have FDA approval.” This tweet misinterprets the “FDA approval of cigarette products,” mistakenly equating the FDA’s marketing authorization with an endorsement of the product’s safety.(23) Another example is the frequent reference to “vape ban” policy. Such claims are exaggerated, as policies do not completely ban the manufacturing and sales of e-cigarettes but rather restrict their reach to youth or ban the sales of flavored products. (32)

#### Distortion/Conspiracy

Distortion/conspiracy claims present a distorted reality and skepticism of established institutions such as the government, businesses, NGOs, and scientific community. These claims typically involve conspiracy narratives about policies, suggesting hidden plots by powerful entities benefiting from a policy at the expense of individual freedom or the general public. A typical example is the claim: “The government bans vape to protect the profits of Big Tobacco/Pharma and can financially benefit from it, at the expenses of small businesses,” or “This is why the #WHO #FCTC fights against vaping, which helps people quit smoking and lowers cancer risk. Tobacco and cancer industries profit $$$.”

### Sources

Finally, we examined the sources likely to spread misinformation. Analysis of the user profiles of the selected tweets showed that those most likely to spread misinformation on Twitter were usually individual vaping advocates. Further investigation of their accounts revealed that these users often had a history of posting pro-vaping content, had fewer followers (Oehmichen et al, 2019), and many had bios or usernames related to the pro-vaping mission. Other misinformation-spreading sources included unspecified individuals or influencers, who are not posting specifically about tobacco-related topics, vaping advocacy groups, and retail stores selling vaping or cannabis products. Individual vaping advocates constituted the largest category of sources, responsible for 79 out of the 169 misinformation tweets, followed by unspecified individuals or influencers (45), vaping advocacy groups (12), and retail stores (12). 31 original tweets were missing, and the sources could not be identified.

While our data collection started with industry tweets, we have not seen misinformation tweets that came from the tobacco industry or about the industry. Instead, in the pro-vaping advocacy tweets, the “Big Tobacco” was seen to be working with the government in the anti-vaping policies, as these policies were seen to be serving their interests to destroy small businesses and individual freedom.

## Discussion

This examination of Twitter data informed the development of an analytical framework for tobacco-related misinformation with three core components: content, type of falsehood, and source. Our framework integrates multiple elements of tobacco-related misinformation. Systematically developing a tobacco-related misinformation framework from social media content with the assistance of generative AI revealed several important characteristics of tobacco-related misinformation and the challenges in identifying and categorizing it.

First, we observed that tobacco-related misinformation often manifests subtly compared with other areas like politics where fabrication and engineered falsehoods are more common (33). Our sample did not see many completely unsupported claims, totally fabricated stories, or “explicit misinformation” (34). In most cases, misinformation typically involved overstated safety or risks, misinterpretation of policies, or misuse of scientific or expert consensus, which has been characterized as “implicit misinformation”.(34) The claims are misleading and ambiguous rather than totally deceptive. This also means that in most cases, the falseness of a tobacco-related claim may not be readily apparent to a non-expert audience.

These nuances in the content may have affected the results of AI selection. As the results showed, only about 1% of the tweets in the sample of 600 tweets (S1) were identified by both human researchers and AI as misinformation, and for S2 (8427 tweets), only 21.7% of the AI-identified tweets (121 out of 557) were valid when assessed by human coders. Most of those tweets had extreme terms and included a distortion of reality or conspiracy attacking governmental agencies or other reputable institutions. AI may have recognized these tweets as misinformation because of the presence of extreme terms, but AI had more difficulty accurately identifying tweets with relative safety claims.

The contentious nature of the discussions and lack of scientific consensus related to new tobacco products made it challenging to categorize claims as misinformation. The analysis of tweets showed that opinions around the safety and risks of e-cigarettes were highly contentious. While neither established guidelines from health authorities like WHO and CDC support e-cigarettes as an effective cessation tool, other health authorities such as the Royal College of Physicians have endorsed e-cigarettes as significantly less harmful and an effective cessation method (30). This divergence in views makes categorizing misinformation and fact-checking difficult and can introduce bias. As these products are relatively new to the market and research may not be able to keep up with the rapid evolution of the tobacco marketplace (6), the lack of up-to-date evidence also contributes to this challenge.

The subtlety of tobacco misinformation also makes equitable information access more difficult. It is more challenging for people with lower levels of information literacy and lack of access to credible sources to identify the subtlety of falseness in claims, making them more susceptible to be misled (2). Researchers, and those designing health campaigns, and interventions should consider addressing ambiguity in messages, and elements in addition to content issues, such as conspiracy theories or source credibility, that may help to identify misinformation.

Second, our findings suggested that misconceptions about policy and regulation are significant misinformation topics, similar to previous research about other health topics that made a distinction between professional (health-related) and political (policy-related) misinformation (35). In our framework, policy was a distinctive misinformation topic with many tweets misinterpreting policies. Such misinterpretations are often part of a distorted view of established authorities, which are seen as serving the interests of the tobacco industry by “banning” vapes, reflecting an overall distrust of institutions (8) that could hinder policies benefiting public health.

The common presence of policy misinformation in the realm of tobacco also suggests that tobacco-related misinformation aligns with a larger network of conspiracy theories in other health topics, such as fluoride (35), vaccine (36), and 5G and COVID-19 (37). These patterned claims with a wide range of variety of topics showed that rather than considering conspiracy theories as a content or theme category as it has been classified in prior studies (14,38,39), it should be a category of falsehood to highlight how this kind of message presents distorted views of reality and makes artificial connections between unrelated facts to foster anti-science and anti-institution agendas.

Additionally, our analysis highlighted that the most common source of tobacco-related misinformation was pro-vaping individuals who did not assume an influencer role, distinct from social bots (40), pro-vaping advocacy groups (10) and influencers (41), which have been previously identified as the misinformation sources in the literature. These findings underscore the need for further investigation, possibly through network analysis, to assess and characterize industry and advocacy group connections to these individuals. Among other sources, pro-vaping advocacy groups on Twitter were known for their presence and the role of “gatekeeping” in spreading misinformation. (10) Retailers could promote products with unsubstantiated health or safety claims that were not authorized for marketing (23), and their roles in spreading misinformation should be further examined.

The development of our framework was supported by a combination of machine learning and human coding to discern subtle nuances in social media posts, along with the use of generative AI for selection and categorization. Although generative AI showed promise in organizing information and aligning with established references, we noted limitations in its keyword matching approach, suggesting the need for iterative prompt adjustments for improved accuracy and more advanced NLP techniques and to explore strategies.

This early-stage exploration, which serves as a first step for formal taxonomy building (15), revealed the complexity of categorizing claims due to contentious discussions and a lack of scientific consensus, particularly concerning new tobacco products like e-cigarettes. Future research should refine and validate our framework by incorporating other media sources, expert opinions (15), and confirmed misinformation content. Future research should also address enhancing the accuracy and utility of AI tools in misinformation detection and categorization.

### Limitations

As an exploratory study, we developed this framework solely based on the textual content of Twitter, which has its own biases in user profiles and language styles. Future research should include other platforms for a diversity of linguistic and media communication features and user characteristics. This preliminary analytical framework needs further development and validation, which could eventually enhance the ability of computer-assisted models or AI to effectively identify tobacco-related misinformation. Another limitation is data availability. This study was limited to tweets from 2020 to 2022. As Twitter became X and the API policy changed, free access for research purpose is no longer possible. There is not currently a viable solution for collecting more data. Additionally, some tweets may have been removed, which may be due to earlier fact-checking efforts made by Twitter.

This study is an exploratory step for further investigation into the utility of generative AI in automated screening for tobacco-related misinformation. The keyword search approach may help flagging certain kind of misinformation posts, such as conspiracy theories and the use of absolute terms on vaping safety. However, AI could not detect the nuances in more complicated claims.

## Conclusion

This study introduces an analytical framework for categorizing tobacco-related misinformation on social media developed through a combination of human expertise and AI-assisted analysis. It highlights the subtle nature of tobacco-related misinformation, which poses significant challenges for non-expert users and individuals with limited information literacy. Given the subtlety and complexity of misinformation on social media, there is a critical need for public health initiatives to communicate clear and precise scientific information to debunk misleading claims. Targeted educational programs should consider these nuances of misinformation to craft tailored messages to guide people, particularly those with lower information literacy and higher rates of tobacco use, on how to access credible sources and effectively utilize information resources. The insights gained from this study can guide public health policymakers and regulatory agencies in effective public communications about new policies and regulatory decisions, taking into account potential misinterpretations and the pervasive spread of misinformation on social media. For example, it may facilitate health communicators to be more proactive in anticipating potential types misinformation and prepare FAQs, evidence-based infographics or other media in advance of releasing new policies.

## Data Availability

All data produced are available online at X.com

